# Automated stratification of trauma injury severity across multiple body regions using multi-modal, multi-class machine learning models

**DOI:** 10.1101/2024.01.22.24301489

**Authors:** Jifan Gao, Guanhua Chen, Ann P. O’Rourke, John Caskey, Kyle Carey, Madeline Oguss, Anne Stey, Dmitriy Dligach, Timothy Miller, Anoop Mayampurath, Matthew M. Churpek, Majid Afshar

**Affiliations:** Department of Biostatistics and Medical Informatics, University of Wisconsin-Madison, Madison, Wisconsin, US; Department of Surgery, University of Wisconsin-Madison, Madison, Wisconsin, US; Department of Medicine, University of Wisconsin-Madison, Madison, Wisconsin, USA; Department of Surgery, Northwestern University Feinberg School of Medicine, Chicago, Illinois, USA; Center of Health Services and Outcomes Research, Institute for Public Health and Medicine, Chicago, Illinois, USA; Department of Computer Science, Loyola University Chicago, Chicago, Illinois, USA; Computational Health Informatics Program, Boston Children’s Hospital, Boston, Massachusetts, United States; Department of Pediatrics, Harvard Medical School, Boston, Massachusetts, United States

## Abstract

The timely stratification of trauma injury severity can enhance the quality of trauma care but it requires intense manual annotation from certified trauma coders. There is a need to establish an automated tool to identify the severity of trauma injuries across various body regions. We gather trauma registry data from a Level I Trauma Center at the University of Wisconsin-Madison (UW Health) between 2015 and 2019. Our study utilizes clinical documents and structured electronic health records (EHR) variables linked with the trauma registry data to create two machine learning models with different approaches to representing text. The first one fuses concept unique identifiers (CUIs) extracted from free text with structured EHR variables, while the second one integrates free text with structured EHR variables. Both models demonstrate impressive performance in categorizing leg injuries, achieving high accuracy with macro-F1 scores of around 0.8. Additionally, they show considerable accuracy, with macro- F1 scores exceeding 0.6, in assessing injuries in the areas of the chest and head. Temporal validation is conducted to ensure the models’ temporal generalizability. We show in our variable importance analysis that the most important features in the model have strong face validity in determining clinically relevant trauma injuries.

## 1 Introduction

Traumatic injuries are the leading cause of death among individuals aged younger than 45, resulting in over 3.5 million hospital admissions in the United States annually [1]. Trauma registries, which collect comprehensive and systematic information, are pivotal in improving trauma care and its related clinical outcomes, as they elucidate injury patterns and identify areas for improvement [2]. Many quality improvement recommendations and policy changes are derived from trauma registries gathered at the local level and reported to state and national trauma databanks [3]. Early assessment of trauma severity, a critical component in trauma registries, is mandatory and clinically significant for correct triage, treatment planning, and resource management [4]. Trauma injury scores, commonly used to gauge the severity of trauma injuries [5], require the annotation by certified trauma coders who utilize software tools to analyze the EHR of trauma admissions [6], which can be time-consuming and involves intense manual labor. In addition, the scores are usually recorded post-discharge, limiting their utility during the patient’s active care. Furthermore, trauma registries are manually curated at trauma centers, leaving a gap in accessing injury scores at non-trauma centers that also triage trauma cases [2]. Automated solutions designed for stratifying injury scores during the time of care have the potential to bridge these gaps. They not only enable more comprehensive and timely data capture but also enhance scalability across various centers.

Machine learning models can learn patterns from existing data and make predictions on unseen data. Recent years have witnessed a surge in the use of machine learning technologies to improve clinical outcomes. Prior research has attempted to use machine learning to automatically predict Abbreviated Injury Scales (AIS) [7], which is one of the most widely used scores [8, 9]. However, these attempts have focused on a single body region and reduced the ordinal AIS scores to binary outcomes. This limitation highlights the necessity for more sophisticated models that can assess multiple body regions with enhanced granularity, thereby meeting the comprehensive documentation needs of trauma registries.

The clinical text contains valuable information [10] frequently used by trauma coders, including radiology reports, operative and procedural notes, history and physical admission notes, etc. One commonly adopted method for modeling unstructured text is to leverage the popular bidirectional encoder representations from transformers (BERT) language model [11]. The variants within BERT families [12, 13, 14] have demonstrated promising performance in a range of clinical text-related tasks [15, 16, 17] including risk stratification [18, 19, 20]. In practical applications, medical concepts can be extracted from the free text and stored in the format of medical concepts from the dictionaries of the Unified Medical Language System (UMLS), also known as concept unique identifiers (CUIs) [21, 22]. The mapped CUIs provide a standardized way of representing clinical concepts from the unstructured clinical free text that is filled with abbreviations and acronyms. Established methods have utilized these CUIs as input to construct machine learning models for clinical risk stratification [23, 24, 25]. Meanwhile, leveraging structured EHR data, including vital signs, laboratory measurements, and clinical scales, offers the potential for predicting trauma-related outcomes [26]. The integration and comparison of different text representations, e.g., CUIs and free text, in conjunction with structured EHR data for modeling trauma severity, remains an unexplored area.

In this study, we have developed two machine learning models to stratify trauma injuries across multiple body regions into three distinct severity levels. These levels are determined based on the magnitude of the AIS and are categorized as negative, minor/moderate, and serious or greater. The first model (denoted as “CUIs + structured EHR model”) handles text converted into CUIs, while the second (denoted as “free text + structured EHR model”) processes unstructured clinical text in free text format, both combined with structured EHR data for a multi-modal input approach. These models aim to accurately predict AIS scores across multiple body regions. The architectures of our models are shown in Figure 1. We present an analysis of their performance, offer both local and global interpretations, and examine the contribution of each modality to the final output.

**Figure 1:**
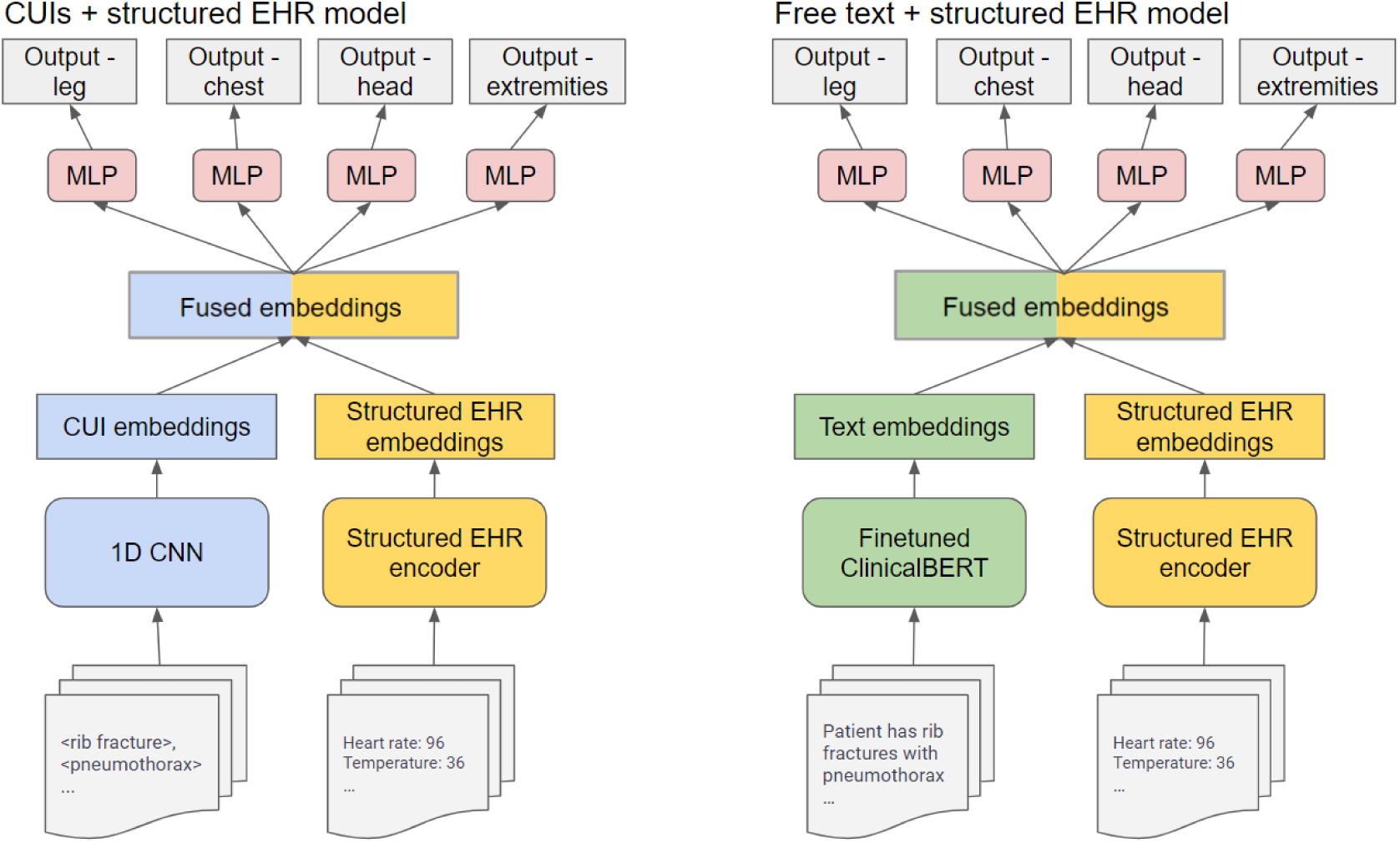
Architecture of the models. We use 1D CNN to encode CUIs and fine-tuned ClinicalBERT to encode free text. A multitask neural network is pretrained on binary severity classification task and its shared layers are leveraged as the encoder for the structured EHR data (see Appendix C). The CUI and free text embeddings, combined with structured EHR data, are fused at an intermediate stage of each model. The concatenated embeddings are then utilized by separated multi-layer perceptrons (MLP) to stratify injury severity across various body regions.

## 2 Methods

### 2.1 Data collection

The cohort used for model development and validation was collected from the UW Health system. Clinical notes and structured EHR data were extracted for trauma patients admitted from 2015 to 2019. Only patients with at least one clinical note available were included in the cohort. The index time was defined as the moment when the first EHR data became available. The data collection window of an admission was defined as 8 hours after the admission. We extracted the ED notes and radiology reports within the data collection window. CUIs were recognized from these notes using cTakes and the engine for mapping the medical concepts from the free text [27]. We also extracted 59 variables that are routinely collected during clinical care from the structured EHR data within the collection window. The descriptions of the structured EHR variables are appended in Appendix A.

### 2.2 Label construction

The AIS used in our dataset was annotated by certified human experts who are trained as trauma registrar coders. The distributions of the original AIS across the nine body regions are presented in Appendix B. Considering that certain categories contain very limited sample sizes in the test set, the nine body regions were collapsed into four combined regions. This categorization was informed by clinical expertise to ensure relevance and accuracy. For each patient:

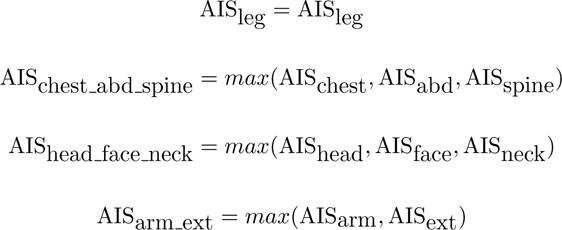

### 2.3 Multimodal data encoding

The CUIs + structured EHR model encoded the CUIs with a one-dimensional convolutional neural network model (1D CNN) [28] and the free text + structured EHR model encoded the free text with a fine-tuned ClinicalBERT [13]. Both models used a pretrained fully-connected neural network to encode the structured EHR variables.

As BERT families have an input length limit, we picked notes that were more likely to offer comprehensive patient information. The ED notes were sorted from long to short and the radiology reports were sorted from early to late. We allocated the first 300 tokens for the sorted ED notes, with the remaining tokens dedicated to the radiology reports. If one of the two note types exceeded its token allocation and the other remained under the limit, we reallocated the unused tokens to accommodate the longer notes. Otherwise, notes that exceeded the limit were truncated accordingly. Following this process, we used the pretrained ClinicalBERT to encode clinical text for each admission.

We used a one-dimensional convolutional neural network model (1D CNN) as the encoder for CUIs. 1D CNN can handle long sequential data and has less complexity compared to other architectures such as transformers and recurrent neural networks (RNN). All the CUIs extracted from the clinical text within the data collection window were encoded by the 1D CNN, without additional need for truncation as the transformer models.

We built a pretrained AIS classifier to encode structured EHR data. As using structured EHR alone to predict the three class severity cannot achieve macro-F1 scores over 0.5, we developed a multitask classifier to predict the binary severity of trauma across various regions with the structured EHR data. The architecture of the binary multitask model is shown in Appendix C. The binary classifier achieved decent discriminative performance and the AUROC were plotted in Appendix D. Then the last shared encoding layer was used as the encoder for structured EHR data in the multiclass task.

### 2.4 Model development

As is shown in Figure 1, the text modality (free text or CUIs) and the structured EHR data were encoded by their encoders and then fused at an intermediate level. After the fusion, the concatenated encoding vectors were fed into four prediction heads. Gradients were passed to the encoders to allow fine-tuning during the training process. To tune the hyperparameters, encounters collected from 2015 to 2017 were for training, and encounters in 2018 were used for tuning. Bayesian optimization [29] was leveraged for efficiently finding optimal hyperparameters. The final models were trained using the tuned hyperparameters on the data collected from 2015 to 2018. A hold-out, temporal test set from 2019 was used to report our results.

### 2.5 Model interpretation

To interpret the output of the models, we computed the attribution scores by leveraging the IG methods. For the free text + structured EHR model, the attribution of each token and structured EHR variable was calculated. Due to the additive property of the IG methods, attribution scores for tokens split from a single word can be aggregated to represent the attribution score of the entire word. For visualizing the risk predictors in individual cases, we assigned varying background colors to words/CUIs and structured EHR variables based on their attribution scores. To achieve a global interpretation of the models, we computed the mean attribution values across all words/CUIs/structured EHR variables.

## 3 Results

### 3.1 Cohort characteristics and outcomes

A total of 9,686 trauma admissions during a five-year study period between 2015 and 2019 were examined. The admissions between 2015 and 2018 (n=7,693) were used for training and tuning, while the admissions in 2019 (n=1,983) served as the test set for temporal validation. We used Abbreviated Injury Scales (AIS) coded by certified human trauma registrar coders. [7]. AIS is a ubiquitous score for trauma injury and ranges from zero to six across nine body regions, with higher scores associated with more critical trauma injuries. The AIS score ground truth labels were categorized into three groups negative (AIS= 0), minor/moderate (0 *<* AIS *<* 3), and serious or greater (AIS *>* 2). Additionally, the body regions were further collapsed into four combined regions according to anatomical properties and with expert annotation by a trauma surgeon (AS): 1. leg; 2. chest, abdomen, and spine (denoted as chest abd spine); 3. head, face, and neck (denoted as head face neck); 4. arm and extremities (denoted as arm ext). The categorized AIS after collapsing are served as the ground truth labels for the supervised training of the models. The cohort characteristics and label distributions are detailed in Table 1.

**Table 1:** Cohort characteristics and label distributions.

|  | Development | Test |
| --- | --- | --- |
| Encounters, n | 7,693 | 1,983 |
| Age, median (IQR) | 59 (38,75) | 63 (44,77) |
| Female, n (%) | 3,225 (41.9) | 855 (43.1) |
| Race |  |  |
| White/Caucasian | 7,096 (92.2) | 1,802 (90.9) |
| Black/African American (%) | 331 (4.3) | 100 (5.0) |
| Asian/Mideast Indian (%) | 102 (1.3) | 34 (1.7) |
| American Indian/Alaska Native (%) | 54 (0.7) | 18 (0.9) |
| Pacific Islander/Hawaiian Native (%) | 18 (0.2) | 1 (< 0.1) |
| Declined/Unknown | 92 (1.1) | 28 (1.4) |
| Death, n (%) | 336 (4.4) | 82 (4.1) |
| Total hours of stay, median (IQR) | 93.9 (53.4,170.1) | 94.7 (51.7,174.3) |
| Number of CUIs, median (IQR) | 188 (113,245) | 192 (122,250) |
| Number of clinical notes, median (IQR) | 14 (10,18) | 13 (9,17) |
| Leg |  |  |
| Negative, n (%) | 4,724 (61.4) | 1,236 (62.3) |
| Minor or moderate, n (%) | 1,466 (19.1) | 339 (17.1) |
| Greater than serious, n (%) | 1,503 (19.5) | 408 (20.6) |
| Chest_abd_spine |  |  |
| Negative, n (%) | 4,345 (56.5) | 1,153 (58.1) |
| Minor or moderate, n (%) | 1350 (17.5) | 329 (16.6) |
| Greater than serious, n (%) | 1,998 (26.0) | 501 (25.3) |
| Head_face_neck |  |  |
| Negative, n (%) | 5,053 (65.7) | 1,351 (68.1) |
| Minor or moderate, n (%) | 1,231 (16.0) | 311 (15.7) |
| Greater than serious, n (%) | 1,409 (18.3) | 321 (16.2) |
| Arm_ext |  |  |
| Negative, n (%) | 2,052 (26.7) | 637 (32.1) |
| Minor or moderate, n (%) | 5,460 (71.0) | 1,303 (65.7) |
| Greater than serious, n (%) | 181 (2.4) | 43 (2.2) |

### 3.2 Model performance

The confusion matrices and the macro-F1 scores scores achieved by the CUIs + structured EHR model and the free text + structured EHR model are displayed in Table 2 and Table 3. The macro-F1 scores scores along with their 95% confidence intervals are represented in Figure 2. The confidence intervals of macro-F1 scores of the models and the corresponding empirical Bayes factors (denoted as *K*) [30] between the two models were computed via bootstrapping with 1, 000 resamplings. A large Bayes Factor (*K >* 10) indicates that one model has significantly superior behavior on the test set compared to the other [31]. The CUIs + structured EHR model model had comparable if not higher macro-F1 scores scores over the free text + structured EHR model on all four body regions. The macro-F1 scores did not significantly differ between the two models except on arm ext with a Bayes Factor larger than 10, where the CUI + structured EHR model achieved nearly 0.6 and the free text + structured EHR model only achieved slightly over 0.5. Both models exhibited their highest performance on leg injury stratification, achieving macro-F1 scores scores of around 0.8. They also demonstrated macro-F1 scores exceeding 0.6 on chest abd spine and head face neck.

**Figure 2:**
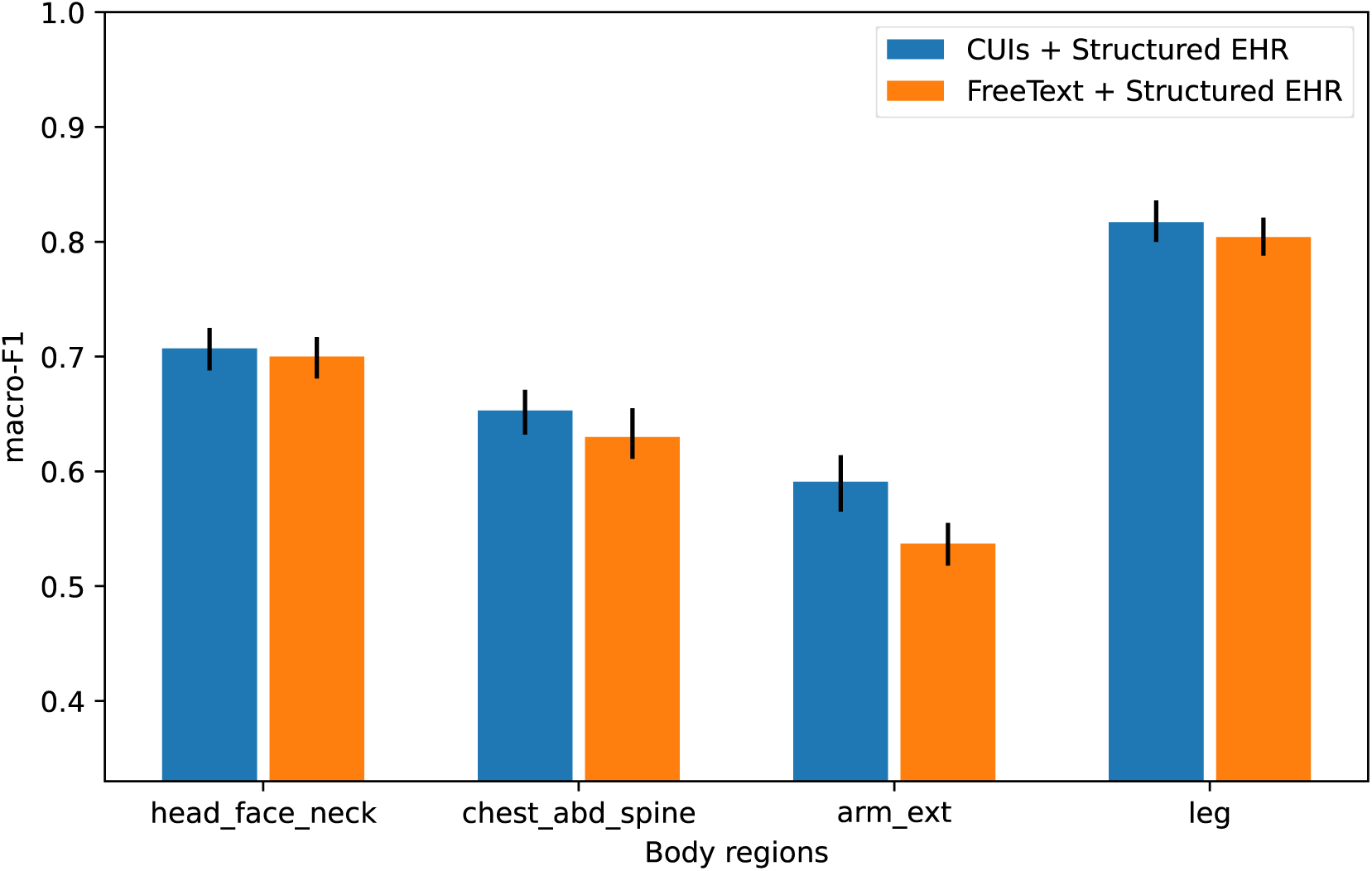
macro-F1 scores and 95% confidence intervals of the two developed models across various body regions. The CUIs + structured EHR model generally achieves macro-F1 scores that are comparable to, or higher than, those of the free text + structured EHR model across all four body regions. Based on Bayes Factors, the difference in performance between the two models is not significant, with the exception of the arm ext region where the CUIs + structured EHR model outperforms the free text + structured EHR model.

**Table 2:**
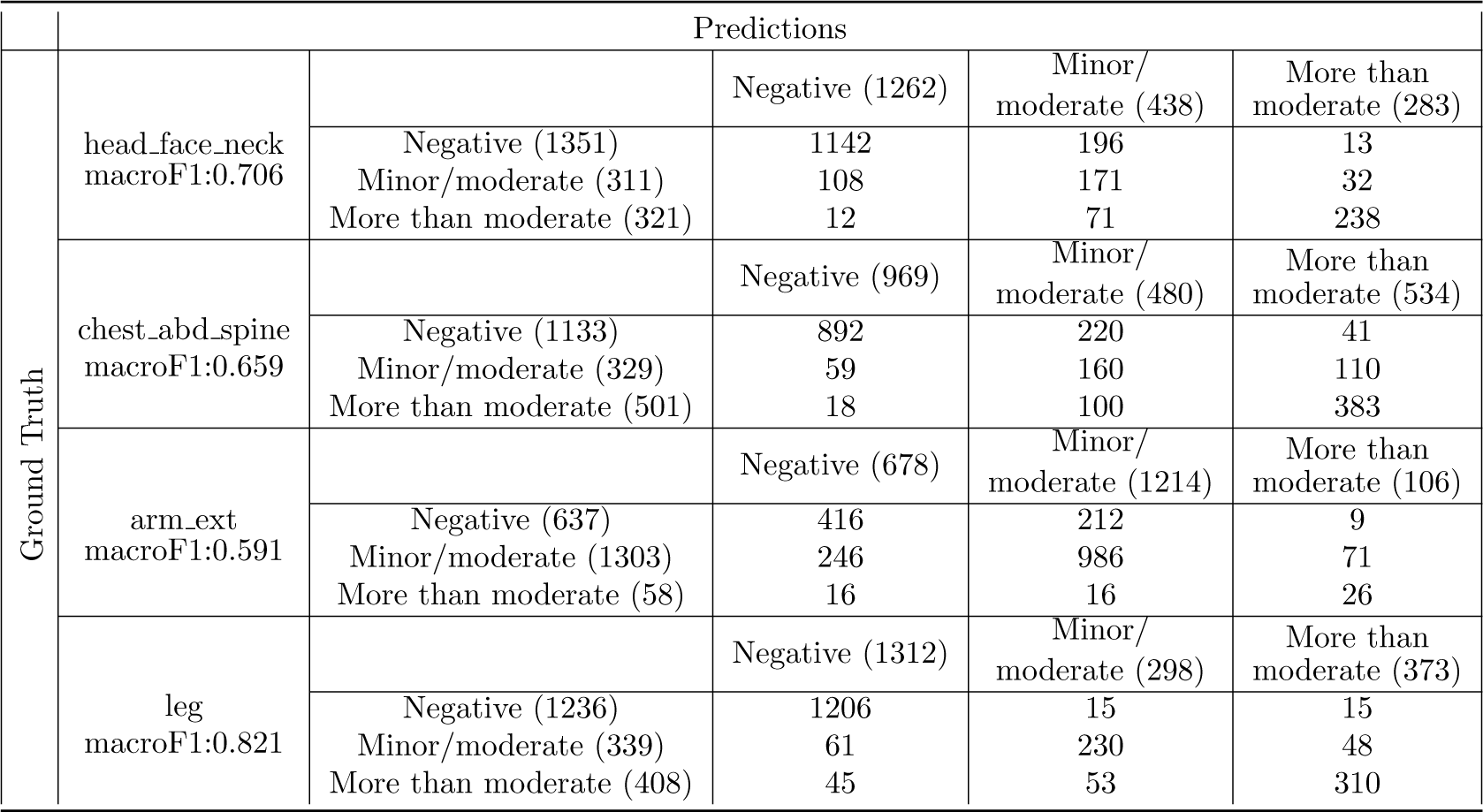
Confusion matrix of the CUIs + structured EHR model. The model excels in stratifying leg injuries with macro-F1 scores near 0.8. It also performs well in assessing injuries in the chest abd spine and head face neck regions, with macro-F1 scores over 0.65.

**Table 3:**
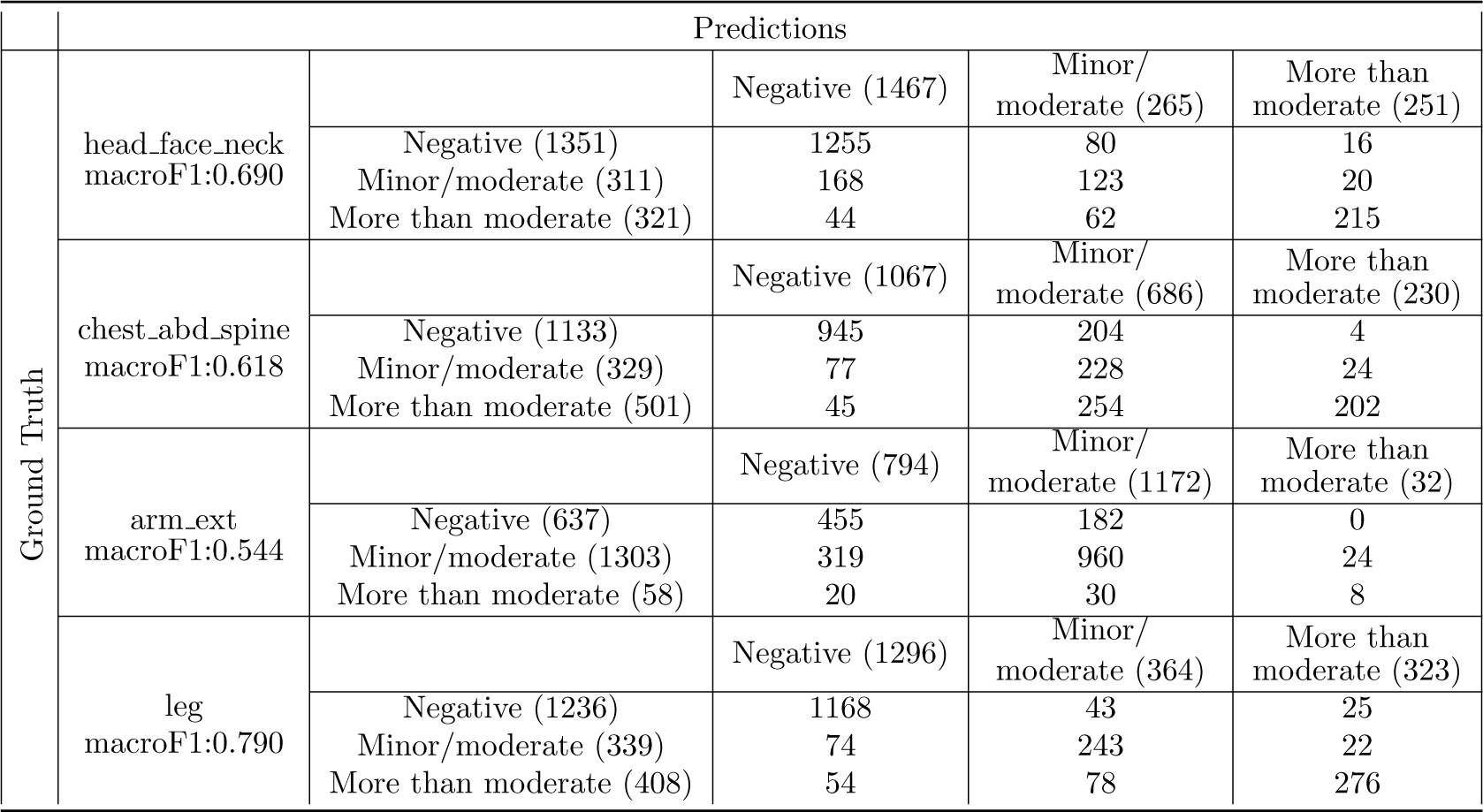
Confusion matrix of the free text + structured EHR model. Similar to the CUIs + structured EHR model, the free text + structured EHR model also achieves excellent performance in leg injury stratification with macro-F1 scores around 0.8. Additionally, it performs well on chest abd spine and head face neck with macro-F1 scores exceeding 0.6.

To further investigate which severity category was more difficult to correctly identify, we evaluated the models’ ability to distinguish each pair of outcomes: Serious vs. Negative, Moderate vs. Negative, Serious vs. Moderate. “Serious” refers to the category of “serious or greater”, while “Moderate” denotes the category of “minor/moderate”. The models’ outputs for each pair of severity categories were compared against the corresponding ground truth labels. The AUROC and the 95% confidence intervals are presented in Figure 3. Both models achieved AUROC over 0.9 when discriminating Serious v.s. Negative samples. However, discriminating the Moderate class with the other two categories proved to be more challenging for both models.

**Figure 3:**
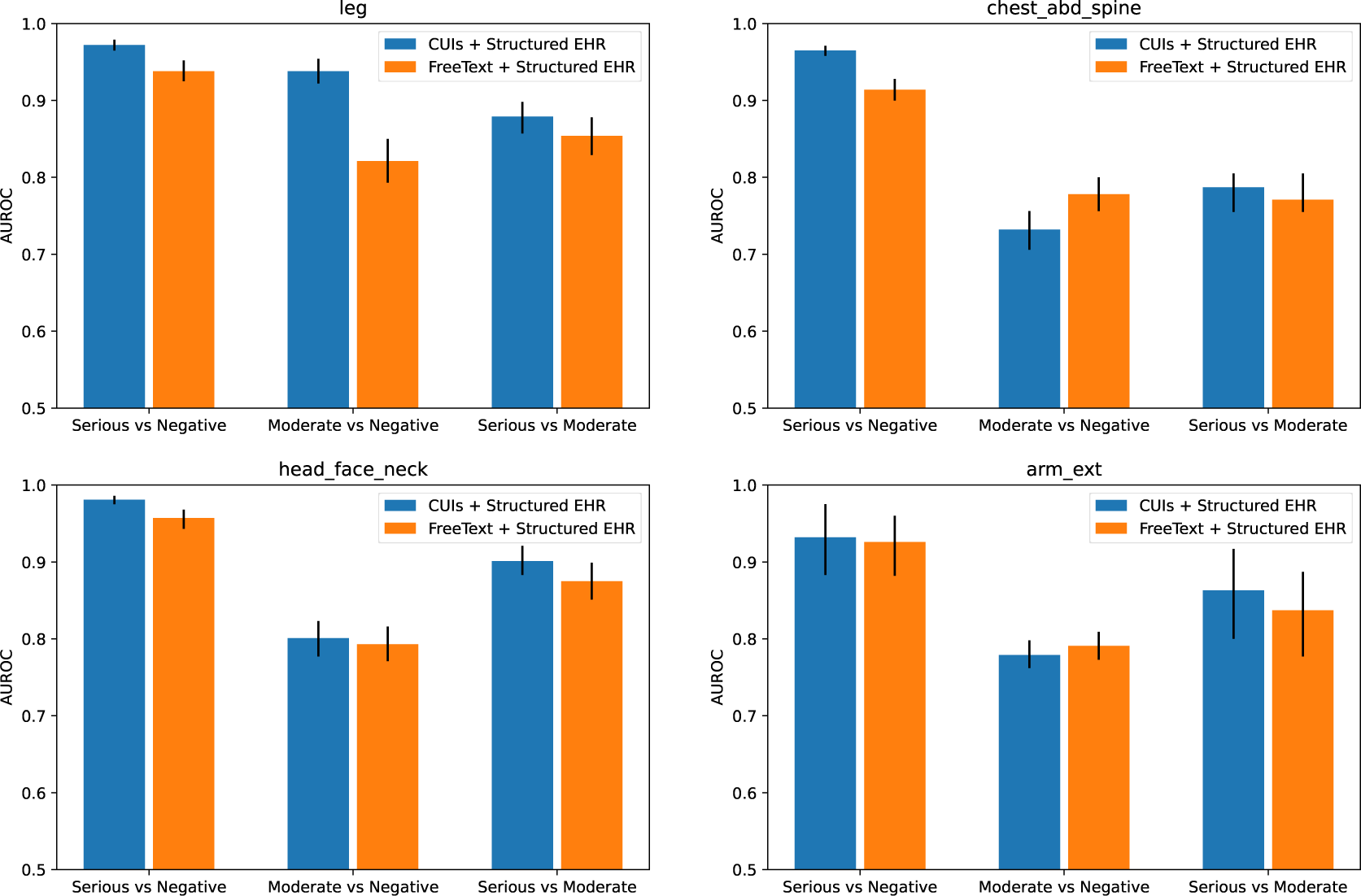
Discrimination performance on pairs of severity levels. Both models demonstrate high accuracy, with AUROC scores exceeding 0.9, in differentiating between Serious and Negative cases. Identifying the Moderate category from the other two is more challenging for both models.

### 3.3 Contribution of structured EHR data

To determine the additive value of structured EHR data in identifying trauma severity, we conducted an ablation study to assess the impact of structured EHR variables on model performance. Table 4 displays the macro F1 scores of our two developed models alongside versions of these models that were built without the structured EHR variables. Based on Bayes Factors, removing structured EHR data did not result in a significant performance decline for either model in three out of the four body regions. The performance only significantly dropped on the arm ext region, which overall was the most challenging region in identifying injury severity according to Figure 2.

**Table 4:** macro-F1 scores and 95% confidence intervals when using different modalities as input. In three of the four body regions, the exclusion of structured EHR data did not lead to a significant performance drop in either model. However, there was a significant decrease in performance in the arm ext region, which was the most challenging area for determining severity in trauma cases according to Figure 2.

|  | CUIs+structured EHR | CUIs | Free text+structured EHR | Free text |
| --- | --- | --- | --- | --- |
| Leg | 0.817 | 0.819 | 0.804 | 0.798 |
|  | (0.800,0.832) | (0.801,0.836) | (0.788,0.821) | (0.780,0.815) |
| Chest_abd.spine | 0.653 | 0.656 | 0.630 | 0.636 |
|  | (0.632,0.671) | (0.638,0.675) | (0.611,0.655) | (0.612,0.659) |
| Head_face.neck | 0.707 | 0.714 | 0.700 | 0.686 |
|  | (0.688,0.725) | (0.694,0.734) | (0.681,0.717) | (0.662,0.707) |
| Arm_ext | 0.591 | 0.552 | 0.537 | 0.499 |
|  | (0.565,0.614) | (0.518,0.585) | (0.518,0.555) | (0.482,0.518) |

We summed the attribution of various modalities via the Integrated Gradient (IG) methods [32] and presented the ratio of attributions of the structured EHR data to the CUIs/free text in Table 5. A higher ratio indicated a more substantial contribution of the structured EHR data to the models’ outputs. The structured EHR data played a more critical role in the CUIs+ structured EHR model compared to the free text + structured EHR model. The structured EHR data had the most crucial impact on arm ext for both models, which was consistent with the results in Table 4 that the arm ext was the only region that significantly benefited from the inclusion of the structured EHR variables.

**Table 5:** Ratio of sum attribution between structured EHR variables and other modalities. The structured EHR data had a more critical role in the CUIs + structured EHR model than in the free text + structured EHR model. Its impact was most pronounced in the arm ext region for both models.

|  | Leg | Chest_abd_spine | Head_face_neck | Arm_ext |
| --- | --- | --- | --- | --- |
| CUIs+structured EHR | 0.215 | 0.417 | 0.371 | 0.929 |
| Free text+structured EHR | 0.051 | 0.069 | 0.055 | 0.101 |

### 3.4 Visualization of cohort-level clinical utility

Figure 4 displays the top 10 CUIs or words with the highest mean attribution scores in the identification of serious or greater injuries determined by the IG methods. Anatomical terms, such as “Bone structure of rib” in CUIs + structured EHR model and “ribs” in the free text + structured EHR model, contributed to the determination of serious cases. Additionally, medical condition concepts such as “Right pneumothorax” in CUIs + structured EHR model and “pneumothorax” in the free text + structured EHR model, were also among the top 10 attributions. The free text + structured EHR model also leveraged descriptive terms such as “displaced” or “rotated” to identify serious cases.

**Figure 4:**
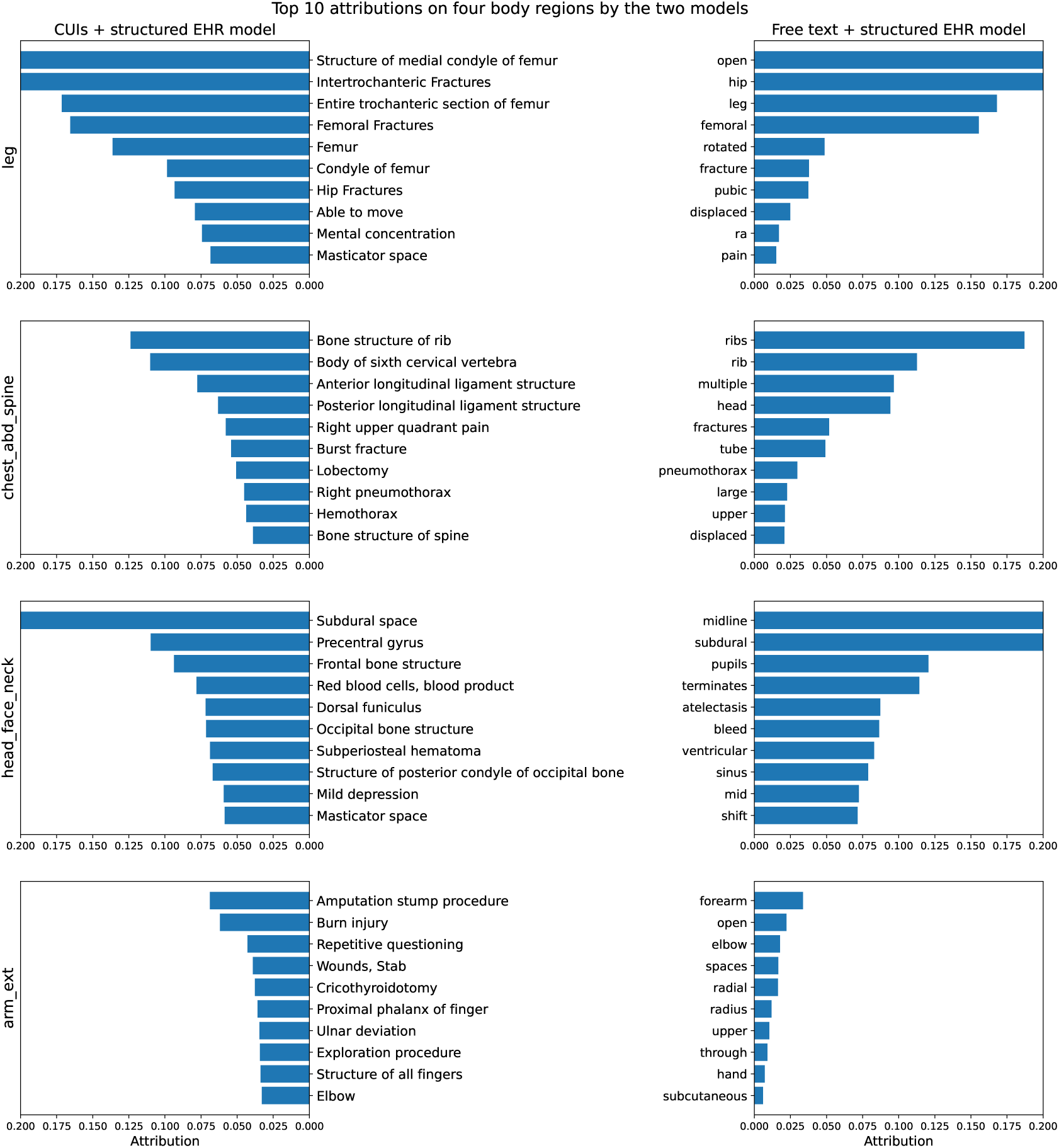
Global level utility. The top 10 attributions are shown for each modal across the four body regions. Attributions over 0.200 are truncated for clean visualizations. Anatomical terms and medical conditions are prominent among the top attributions. The free text + structured EHR model utilizes descriptive terms as well.

### 3.5 Visualization of individual-level clinical utility

To illustrate the clinical utility at the individual level provided by our models, Figure 5 and Figure 6 provide visual examples illustrating how a combination of CUIs or free text, and structured EHR data contributed to identifying a serious or greater chest injury. In these visualizations, CUIs and words were highlighted with warmer background colors to indicate an increased likelihood of serious injuries in the combined chest region, while cooler colors suggested a decreased probability. The top 10 contributing structured EHR variables were also shown in the visualization. The height of each bar in the plot corresponded to the percentile ranking of the structured EHR data and the color temperature reflected its contribution to the likelihood of a serious injury, akin to the color coding used for CUIs and text. The output prediction, baseline score used by the IG method, total attribution of modalities, and the approximation error of the IG algorithm were also presented at the top of the visualizations.

**Figure 5:**
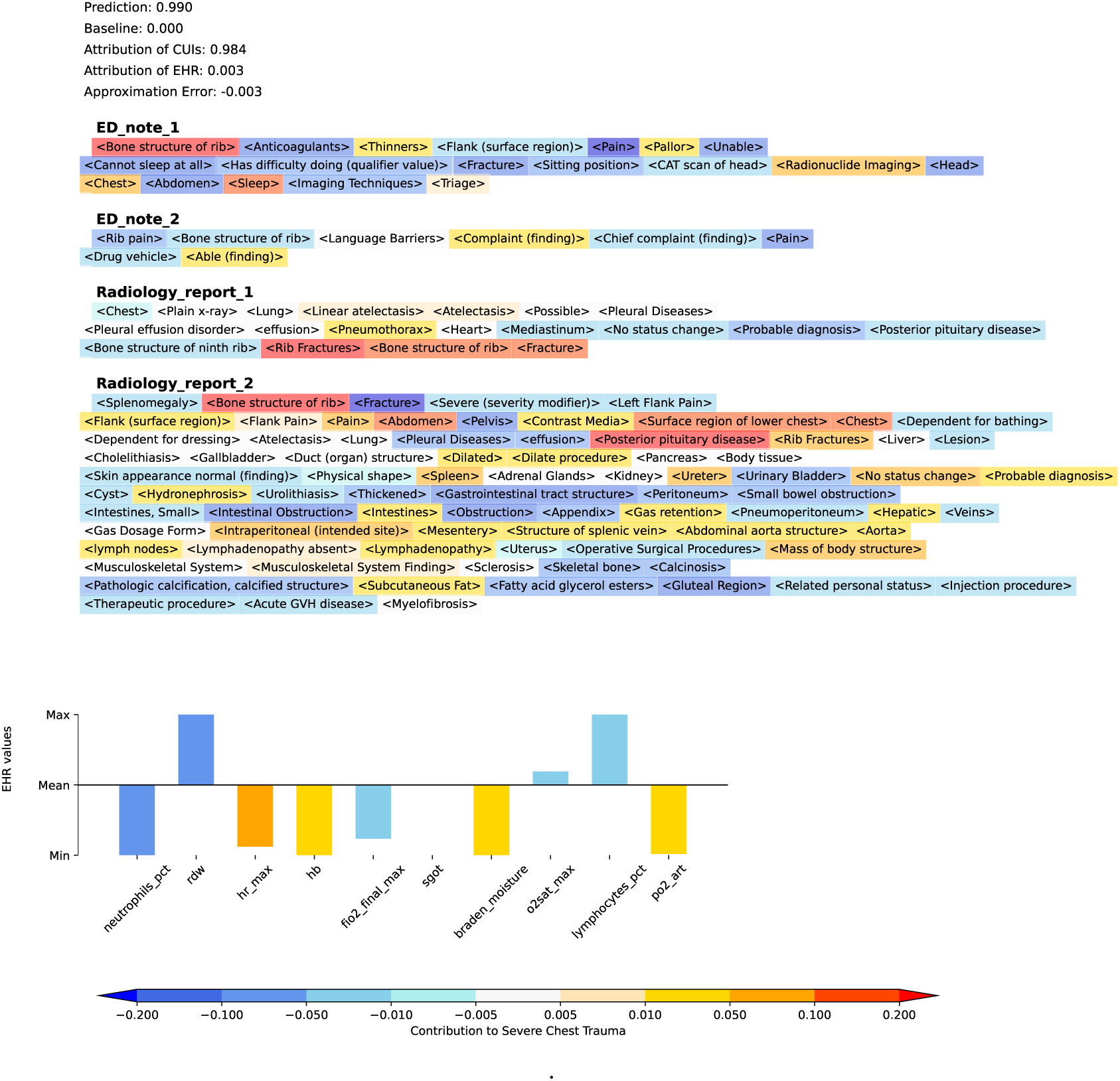
A visualization example of the individual-level clinical utility for the CUIs + structured EHR model. A warmer background color indicated an increased chance of having serious injuries in the combined chest region, while cooler colors reflected a decreased probability. This was the same encounter as Figure 6

**Figure 6:**
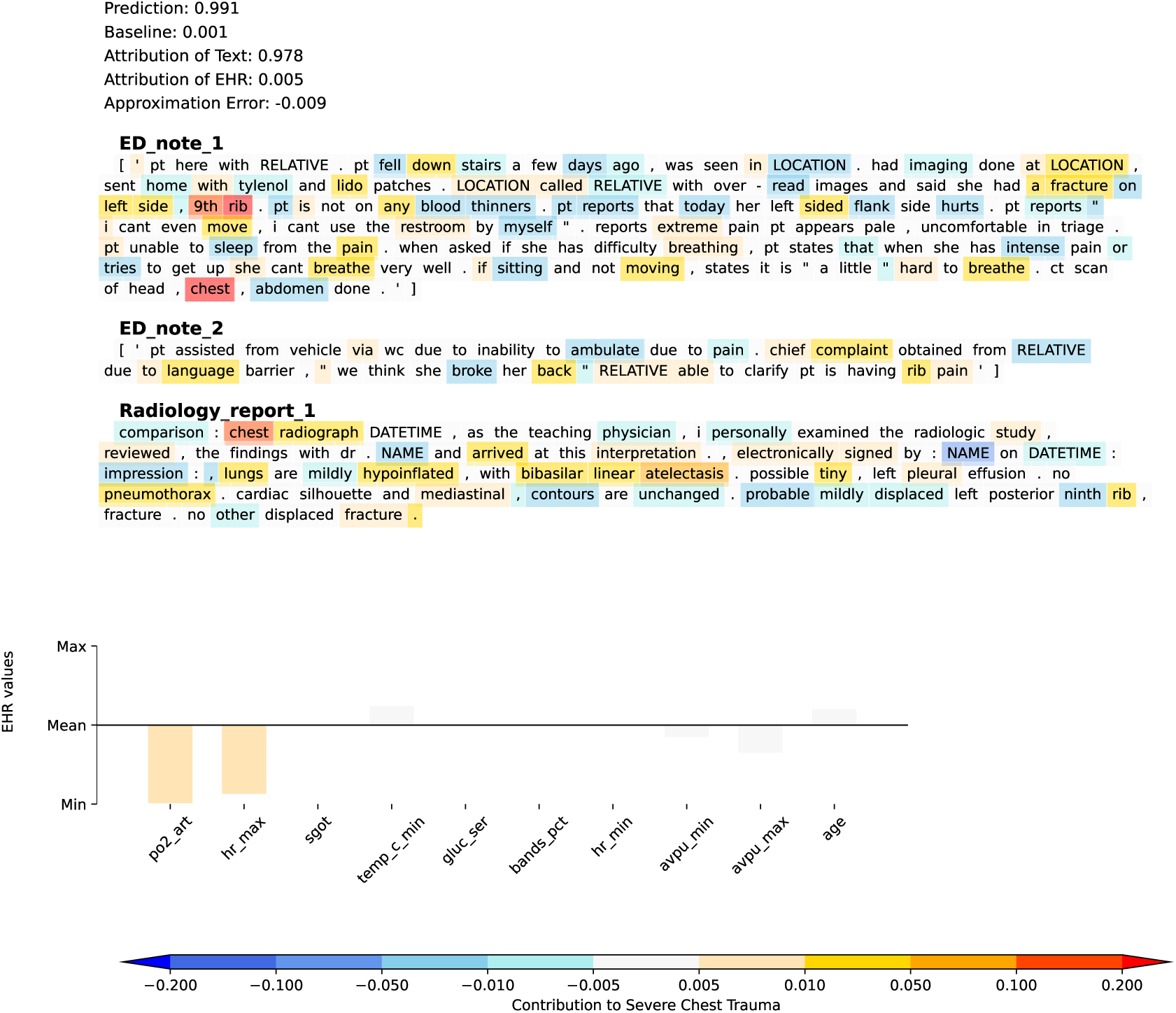
A visualization example of the individual-level clinical utility for the free text + structured EHR model. Akin to Figure 5, a warmer background color indicated the presence of certain words increased the likelihood of having serious injuries in the combined chest region. Conversely, cooler colors indicated that the occurrence of these words was associated with a lower likelihood of serious injuries. This is the same encounter as Figure 5

## 4 Discussion

We developed a multiclass model for the automatic stratification of trauma injury severity using real-world clinical data. To the best of our knowledge, this is the first work that identifies trauma injury severity across multiple body regions beyond binary classification. In practice, our model shows great potential to be implemented in clinical settings capable of collecting clinical text, whether as free text or CUIs, along with structured EHR data.

We used IG methods to capture the attribution of input features to model predictions. Shapley value based methods such as SHAP (SHapely Additive exPlanations)[33] are commonly used for model explanation. However, these methods are typically less suited for unstructured data. In addition, these methods rely on sampling from an exponentially large feature space and permuting all possible feature values whereas the IG methods compute the model’s gradient fewer than 100 times, making them more efficient for clinical applications with differentiable models. Furthermore, the IG methods adhere to the “Implementation invariance” axiom, meaning that the attribution of two models should be identical if their outputs are equal for all inputs. Some popular interpretation methods such as DeepLift [34] and LRP [35] fail to fulfill this requirement. By leveraging IG methods, our models can provide both local interpretation and global interpretation. The global interpretation sheds light on which features (whether word tokens in free text, medical concepts in CUIs, or measurements from structured EHR data) contribute most to the final output. For individual trauma cases, medical staff can gain insights into the rationale behind our models’ decisions through heatmap visualizations, enhancing understanding and trust in the predictions at the point of care.

Though applying BERT-family models to the clinical text has demonstrated advantages in addressing real-world challenges, the limit of input token length and the large number of parameters in the attention mechanism may hinder their application in clinical practice. CUIs from the Unified Medical Language Systems (UMLS) [36] can be extracted from clinical text using tools such as cTakes [27] and MetaMap [37]. The CUI representation focuses more precisely on medical concepts, filtering out non-medical-related elements like stopwords and converting various expressions of medical terms in free text into concepts within standardized vocabularies. Furthermore, compared to modeling free text with BERT-family models, the CUIs can be encoded by models that have fewer parameters and are not constrained by the length of the input. In our study, the CUIs + structured EHR model that leverages 1D CNN to encode the CUIs contains 3,122,709 parameters. In contrast, the free text + structured EHR model that utilizes transformers for encoding free text encompasses a total of 108,372,469 parameters. Meanwhile, the CUIs + structured EHR model achieves comparable, if not superior, performance to the free text + structured EHR model in terms of macro-F1 scores, which indicates that omitting the context information of medical concepts from the clinical text does not impede the ability of machine learning models to discern trauma injury severity. Though extracting CUIs from clinical text requires additional processing steps, these steps are reusable across applications and our health system currently has a real-time CUI-based pipeline in place for other health systems to benchmark [22]. Our research highlights the potential of leveraging CUI-based methods to develop accurate, generalizable, and parameter-efficient models for clinical applications.

Based on Figure 4, both models prioritize anatomical regions such as rib, femur, and subdural spaces to identify the location of trauma injuries. It is consistent with clinical practice because anatomical regions are usually the initial indicators for categorizing body injuries. Both models can also recognize severe medical conditions such as hemothorax as indicators of injury severity. However, one notable difference in the CUIs + structured EHR model is that the concept “Fracture” is not among the top attributions for the combined chest and leg regions, unlike in the free text + structured EHR model where the words “fracture” and “fractures” are primary attributions at these two regions. This discrepancy can be explained by the fact that mentions of “fracture” can pertain to various regions, rendering the term less specific in determining the severity of injuries in a particular area. On the other hand, BERT-family models can contextualize each token, making the word ‘fracture’ a more reliable indicator of injury severity when combined with surrounding semantic information. In addition, the free text + structured EHR model can also capture adjective descriptions such as “displaced” and “rotated” which are usually overlooked by CUIs. Though the difference in model behaviors exists, the numerical performance is comparable across the body regions except in the combined extremities. As our development set has a very limited number of samples for serious or greater injuries on the region of arm and extremities, the heavily parametrized ClinicalBERT may fail to capture sufficient information to be generalized, whereas the less complicated 1D CNN may offer better generalizability when the label distribution is highly skewed.

The findings from Table 4 and 5 reveal that information from the structured EHR data has a crucial impact and can benefit the models for the region of arm and extremities, which is characterized by a skewed label distribution and both models underperform on compared to other body regions. In Figure 4, strong predictors are not discernible from either the free text or the CUIs on arm ext. Such observations indicate that incorporating the structured EHR data can provide additive values in predictive powers when the signals from the clinical text are weak, though such improvements are less pronounced when the text data alone is strongly indicative.

While the AIS is a commonly used metric for classifying the severity of trauma injuries, its primary design is for the evaluation of individual injuries. This focus may limit its ability to capture the cumulative effect of multiple injuries [38]. Our model, which employs AIS as the ground truth for injury severity, may be applied to more comprehensive scores such as the Injury Severity Score [39] that incorporates AIS into an overall trauma severity. Another potential limitation of our study is the absence of external validation for our models. While external validation is generally recommended for locally developed machine learning models, it cannot guarantee the usefulness of the model due to the fact it may not reflect the characteristics of the intended population [40]. As our primary goal is to take the initial step towards assessing the feasibility and significance of integrating various modalities for automatic trauma injury stratification, achieving universal generalizability is not our foremost concern in this study. To ensure the robustness of our models to the dynamic nature of healthcare, we have conducted thorough temporal validation using a recently collected local dataset.

In conclusion, we built multimodal machine learning models using two text representations, CUIs and free text, in conjunction with structured EHR data separately. The CUIs + structured EHR model achieves comparable or superior performance over the free text + structured EHR model in macro-F1 scores across all four body regions. Both models can provide clinically relevant interpretations. Our work also highlights that incorporating structured EHR data into the models helps to mitigate underperformance when the text modality is not strongly indicative.

## Data Availability

All data produced in the present study are available upon reasonable request to the authors.

## Appendix

### A Names and featurization of structured EHR variables

**Table A1:** The first column shows the names of vital signs and lab measurements that are extracted from the structured EHR data. The second column shows the featurization of the structured EHR data variables. “Max & Min” means the max and min values are extracted within the first 8 hours of the encounter and “First” means the first measurements within the first 8 hours of the encounter are used as features.

| Name | Featurization |
| --- | --- |
| hr | Max & Min |
| rr | Max & Min |
| sbp | Max & Min |
| dbp | Max & Min |
| o2sat | Max & Min |
| temp_c | Max & Min |
| avpu | Max & Min |
| bmi | First |
| fio2_final | Max & Min |
| braden_activity | First |
| braden_friction | First |
| braden_mobility | First |
| braden_moisture | First |
| braden_nutrition | First |
| braden_sensory | First |
| braden_scale | First |
| urine_output_sum | First |
| albumin | First |
| alk_phos | First |
| bands_pct | First |
| bili_total | First |
| bun | First |
| calcium | First |
| chloride | First |
| co2 | First |
| creatinine | First |
| eosinophils_pct | First |
| gluc_ser | First |
| hemoglobin | First |
| inr | First |
| lactate | First |
| lipase | First |
| lymphocytes_pct | First |
| magnesium | First |
| mcv | First |
| monocytes_pct | First |
| neutrophils_pct | First |
| pco2_art | First |
| pco2_ven | First |
| ph_art | First |
| ph_ven | First |
| phosphate | First |
| platelet_count | First |
| po2_art | First |
| potassium | First |
| ptt | First |
| rdw | First |
| ast_sgot | First |
| sodium | First |
| total_protein | First |
| wbc | First |
| anion_gap | First |
| timeofday | First |
| age | First |

### B Original AIS distribution

**Figure B1:**
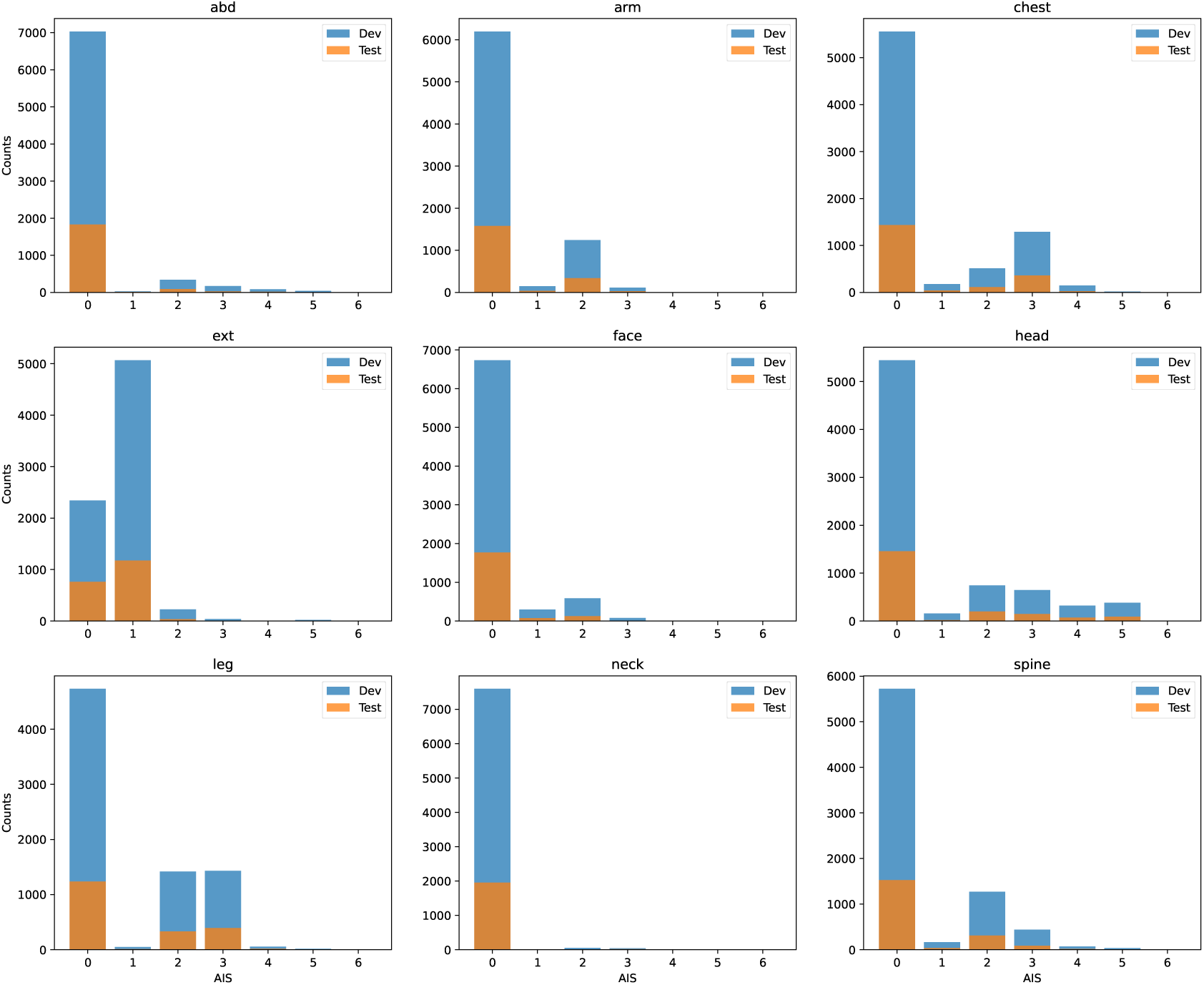
Original AIS across all the nine AIS regions.

### C Construction of the structured EHR encoder

**Figure C1:**
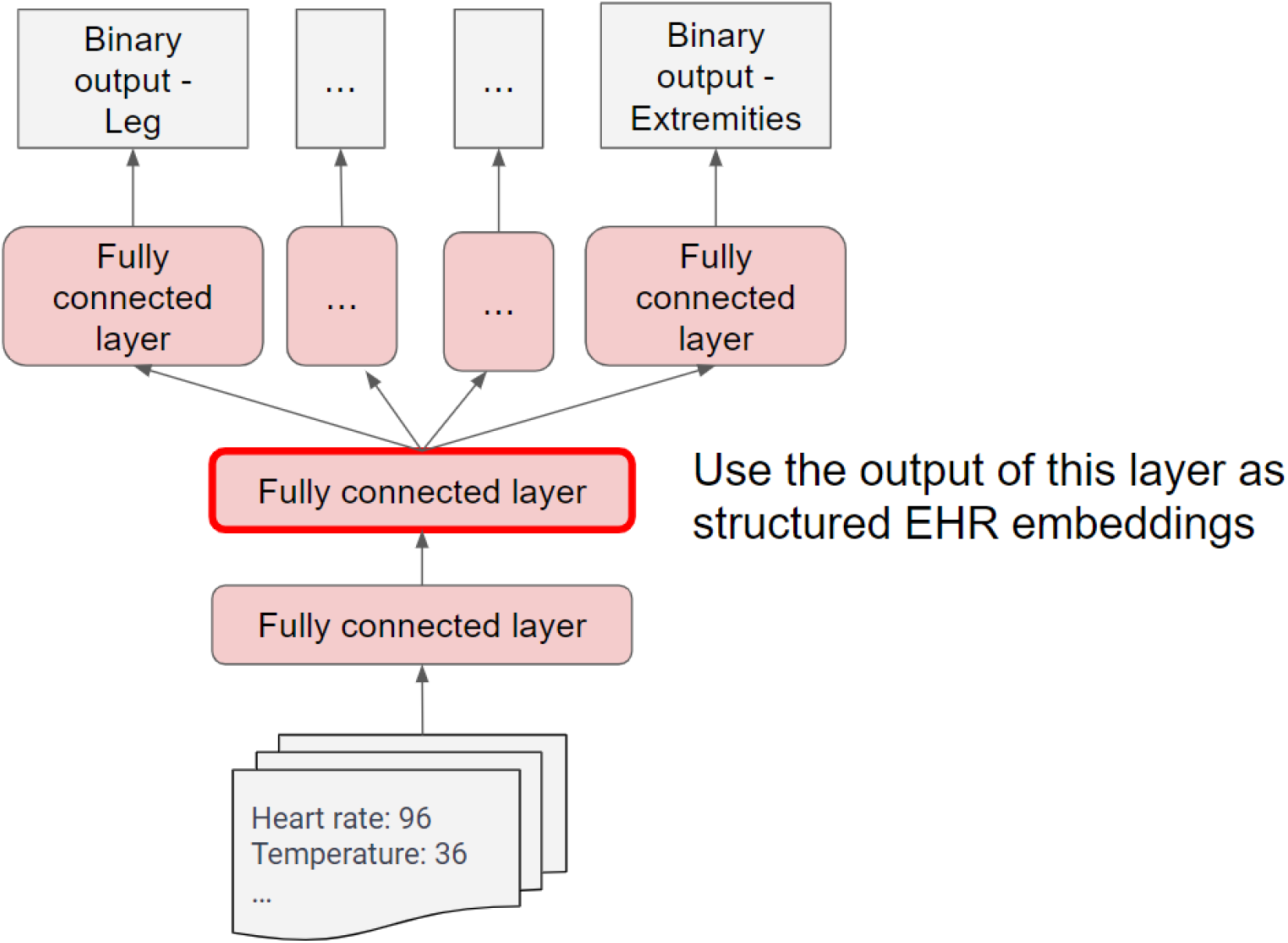
The architecture of the binary multitask model that is utilized for embedding structured EHR data. The shared architecture includes two fully connected layers and the model also contains separated layers for different body regions. The output of the last shared layer can be used as the structured EHR embeddings that provide information on trauma severity.

### D Discriminative performance of the binary structured EHR model

**Figure D1:**
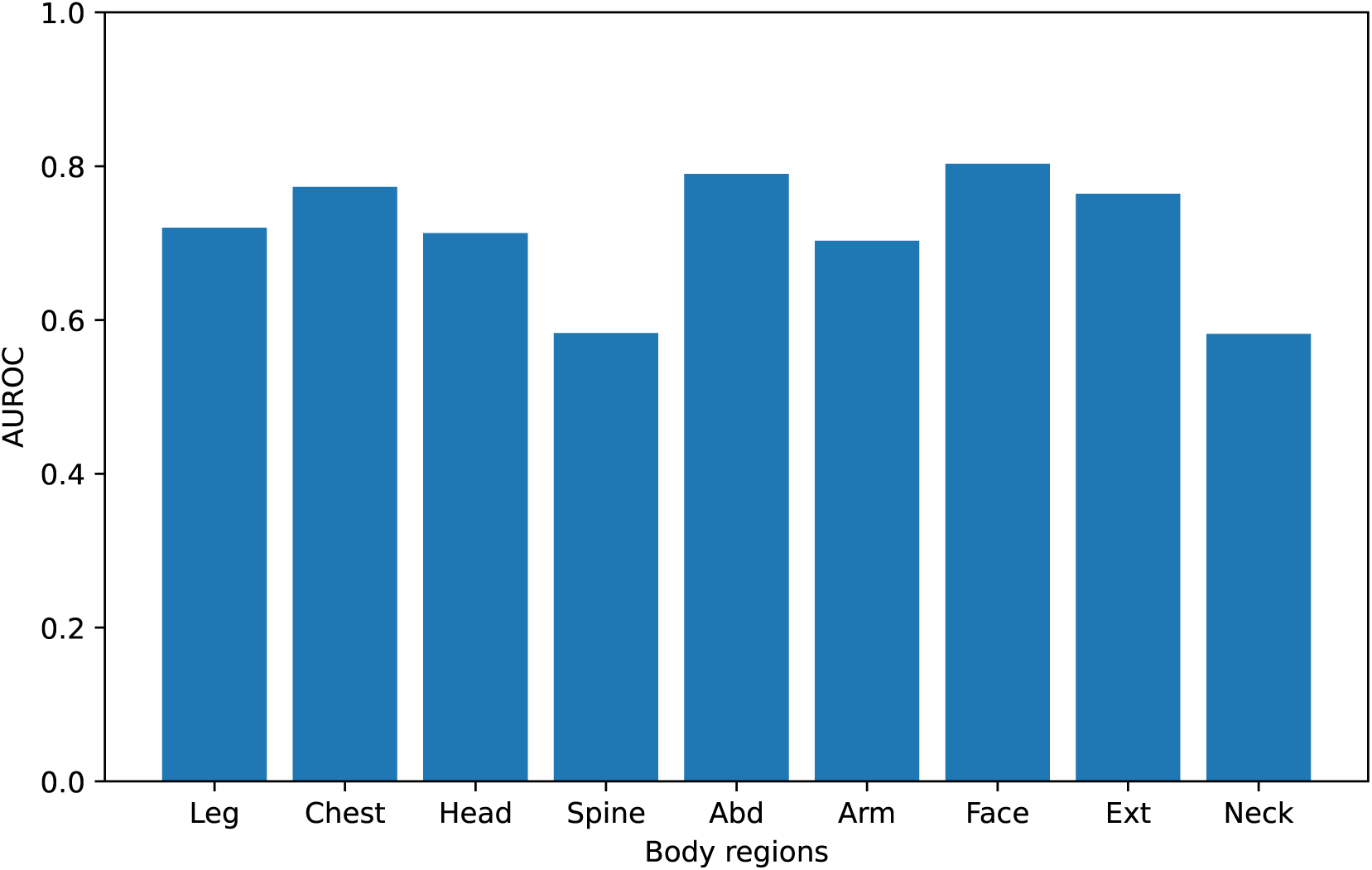
AUROC and AUPR when the pretrained structured EHR model makes binary output across the nine AIS body regions.

## References

[1] Herrera-Escobar JP, Schneider JC. From survival to survivorship—framing traumatic injury as a chronic condition. The New England journal of medicine. 2022;387(7):581.

[2] Moore L, Clark DE. The value of trauma registries. Injury. 2008;39(6):686–95.

[3] Cameron PA, Gabbe BJ, McNeil JJ, Finch CF, Smith KL, Cooper DJ, et al. The trauma registry as a statewide quality improvement tool. Journal of Trauma and Acute Care Surgery. 2005;59(6):1469–76.

[4] Champion HR, Sacco WJ, Hunt TK. Trauma severity scoring to predict mortality. World journal of surgery. 1983;7:4–11.

[5] Mock C. WHO releases Guidelines for trauma quality improvement programmes. BMJ Publishing Group Ltd; 2009.

[6] Palmer CS, Gabbe BJ, Cameron PA. Defining major trauma using the 2008 Abbreviated Injury Scale. Injury. 2016;47(1):109–15.

[7] Friedman Z, Kugel C, Hiss J, Marganit B, Stein M, Shapira SC. The Abbreviated Injury Scale: a valuable tool for forensic documentation of trauma. The American journal of forensic medicine and pathology. 1996;17(3):233–8.

[8] Kulshrestha S, Dligach D, Joyce C, Baker MS, Gonzalez R, O’Rourke AP, et al. Prediction of severe chest injury using natural language processing from the electronic health record. Injury. 2021;52(2):205–12.

[9] Kulshrestha S, Dligach D, Joyce C, Gonzalez R, O’Rourke AP, Glazer JM, et al. Comparison and interpretability of machine learning models to predict severity of chest injury. JAMIA open. 2021;4(1):ooab015.

[10] Seinen TM, Fridgeirsson EA, Ioannou S, Jeannetot D, John LH, Kors JA, et al. Use of unstructured text in prognostic clinical prediction models: a systematic review. Journal of the American Medical Informatics Association. 2022;29(7):1292–302.

[11] Devlin J, Chang MW, Lee K, Toutanova K. Bert: Pre-training of deep bidirectional transformers for language understanding. arXiv preprint arXiv:181004805. 2018.

[12] Rasmy L, Xiang Y, Xie Z, Tao C, Zhi D. Med-BERT: pretrained contextualized embeddings on large-scale structured electronic health records for disease prediction. NPJ digital medicine. 2021;4(1):86.

[13] Alsentzer E, Murphy JR, Boag W, Weng WH, Jin D, Naumann T, et al. Publicly available clinical BERT embeddings. arXiv preprint arXiv:190403323. 2019.

[14] Lee J, Yoon W, Kim S, Kim D, Kim S, So CH, et al. BioBERT: a pre-trained biomedical language representation model for biomedical text mining. Bioinformatics. 2020;36(4):1234–40.

[15] Pawar Y, Henriksson A, Hedberg P, Naucler P. Leveraging clinical bert in multimodal mortality prediction models for covid-19. In: 2022 IEEE 35th International Symposium on Computer-Based Medical Systems (CBMS). IEEE; 2022. p. 199–204.

[16] He Y, Zhu Z, Zhang Y, Chen Q, Caverlee J. Infusing disease knowledge into BERT for health question answering, medical inference and disease name recognition. arXiv preprint arXiv:201003746. 2020.

[17] Gao J, He S, Hu J, Chen G. A hybrid system to understand the relations between assessments and plans in progress notes. Journal of Biomedical Informatics. 2023;141:104363.

[18] Alambo A, Andrew R, Gollarahalli S, Vaughn J, Banerjee T, Thirunarayan K, et al. Measuring pain in sickle cell disease using clinical text. In: 2020 42nd Annual International Conference of the IEEE Engineering in Medicine & Biology Society (EMBC). IEEE; 2020. p. 5838–41.

[19] Fanconi C, van Buchem M, Hernandez-Boussard T. Natural Language Processing Methods to Identify Oncology Patients at High Risk for Acute Care with Clinical Notes. AMIA Summits on Translational Science Proceedings. 2023;2023:138.

[20] Islam MS, Rahman W, Abdelkader A, Yang PT, Lee S, Adams JL, et al. Using AI to Measure Parkinson’s Disease Severity at Home. arXiv preprint arXiv:230317573. 2023.

[21] Afshar M, Dligach D, Sharma B, Cai X, Boyda J, Birch S, et al. Development and application of a high throughput natural language processing architecture to convert all clinical documents in a clinical data warehouse into standardized medical vocabularies. Journal of the American Medical Informatics Association. 2019;26(11):1364–9.

[22] Afshar M, Adelaine S, Resnik F, Mundt MP, Long J, Leaf M, et al. Deployment of Real-time Natural Language Processing and Deep Learning Clinical Decision Support in the Electronic Health Record: Pipeline Implementation for an Opioid Misuse Screener in Hospitalized Adults. JMIR Medical Informatics. 2023;11:e44977.

[23] Gong JJ, Naumann T, Szolovits P, Guttag JV. Predicting clinical outcomes across changing electronic health record systems. In: Proceedings of the 23rd ACM SIGKDD international conference on knowledge discovery and data mining; 2017. p. 1497–505.

[24] Ye J, Yao L, Shen J, Janarthanam R, Luo Y. Predicting mortality in critically ill patients with diabetes using machine learning and clinical notes. BMC medical informatics and decision making. 2020;20(11):1–7.

[25] Liang JJ, Lehman E, Iyengar A, Mahajan D, Raghavan P, Chang CY, et al. Towards generalizable methods for automating risk score calculation. In: Proceedings of the 21st Workshop on Biomedical Language Processing; 2022. p. 426–31.

[26] Zhang T, Nikouline A, Lightfoot D, Nolan B. Machine learning in the prediction of trauma outcomes: A systematic review. Annals of emergency medicine. 2022.

[27] Savova GK, Masanz JJ, Ogren PV, Zheng J, Sohn S, Kipper-Schuler KC, et al. Mayo clinical Text Analysis and Knowledge Extraction System (cTAKES): architecture, component evaluation and applications. Journal of the American Medical Informatics Association. 2010;17(5):507–13.

[28] Alzubaidi L, Zhang J, Humaidi AJ, Al-Dujaili A, Duan Y, Al-Shamma O, et al. Review of deep learning: Concepts, CNN architectures, challenges, applications, future directions. Journal of big Data. 2021;8:1–74.

[29] Turner R, Eriksson D, McCourt M, Kiili J, Laaksonen E, Xu Z, et al. Bayesian optimization is superior to random search for machine learning hyperparameter tuning: Analysis of the black-box optimization challenge 2020. In: NeurIPS 2020 Competition and Demonstration Track. PMLR; 2021. p. 3–26.

[30] Bergquist T, Schaffter T, Yan Y, Yu T, Prosser J, Gao J, et al. Evaluation of crowd-sourced mortality prediction models as a framework for assessing AI in medicine. medRxiv. 2021:2021–01.

[31] Kass RE, Raftery AE. Bayes factors. Journal of the american statistical association. 1995;90(430):773–95.

[32] Sundararajan M, Taly A, Yan Q. Axiomatic attribution for deep networks. In: International conference on machine learning. PMLR; 2017. p. 3319–28.

[33] Lundberg SM, Lee SI. A unified approach to interpreting model predictions. Advances in neural information processing systems. 2017;30.

[34] Shrikumar A, Greenside P, Kundaje A. Learning important features through propagating activation differences. In: International conference on machine learning. PMLR; 2017. p. 3145–53.

[35] Binder A, Montavon G, Lapuschkin S, Müller KR, Samek W. Layer-wise relevance propagation for neural networks with local renormalization layers. In: Artificial Neural Networks and Machine Learning–ICANN 2016: 25th International Conference on Artificial Neural Networks, Barcelona, Spain, September 6-9, 2016, Proceedings, Part II 25. Springer; 2016. p. 63–71.

[36] Bodenreider O. The unified medical language system (UMLS): integrating biomedical terminology. Nucleic acids research. 2004;32(suppl 1):D267–70.

[37] Aronson AR. Metamap: Mapping text to the umls metathesaurus. Bethesda, MD: NLM, NIH, DHHS. 2006;1:26.

[38] Asensio JA, Trunkey DD. Current Therapy of Trauma and Surgical Critical Care E-Book. Elsevier Health Sciences; 2008.

[39] Baker SP, o’Neill B, Haddon Jr W, Long WB. The injury severity score: a method for describing patients with multiple injuries and evaluating emergency care. Journal of Trauma and Acute Care Surgery. 1974;14(3):187–96.

[40] Youssef A, Pencina M, Thakur A, Zhu T, Clifton D, Shah NH. External validation of AI models in health should be replaced with recurring local validation. Nature Medicine. 2023:1–2.

